# From Power Spectral Density to Wavelets: Improving Symbolic Representations of Electroencephalography Band Dynamics in the Weed Plot Framework

**DOI:** 10.64898/2026.05.05.26352441

**Authors:** Vanesa B. Meinardi, Carina Boyallian, Rocio S. Giuzio

## Abstract

Electroencephalography (EEG) interpretation in clinical practice relies on the analysis of energy distribution across standard frequency bands. The Weed Plot framework encodes band-wise spectral energy, computed using Fourier-based methods, into a symbolic representation that preserves the interpretability of traditional EEG analysis.

In this study, we propose a wavelet-based extension of this framework, where the energy of predefined clinical EEG bands is estimated using the Discrete Wavelet Transform instead of Power Spectral Density. Unlike Fourier-based approaches, wavelets provide a time–frequency representation that captures transient and non-stationary dynamics while remaining consistent with clinically defined bands.

From these estimates, symbolic patterns are constructed based on the relative ordering of frequency bands within short temporal windows. Their empirical distribution is used to extract entropy-based features for epilepsy detection using multiple machine learning classifiers.

From an Artificial Intelligence perspective, the main contribution is a structured symbolic encoding that enhances feature discriminability. From an engineering perspective, the contribution lies in an automated framework for EEG-based epilepsy detection. Experimental results show that wavelet-based representations improve classification performance compared to raw entropy and Fourier-based features. This improvement arises from the interaction between time–frequency localization and symbolic encoding, producing more discriminative feature distributions.

These findings support wavelet-based symbolic representations as a robust and interpretable framework for EEG analysis, bridging clinical interpretation and data-driven methods.

## 1. Introduction

Electroencephalography (EEG) interpretation in clinical practice relies on analyzing the distribution of signal energy across standard frequency bands [7, 26]. By recording synaptic activity through scalp electrodes, EEG provides a non-invasive tool for the diagnosis and monitoring of neurological conditions such as epilepsy, sleep disorders, and encephalopathies [33]. Although widely used, clinical EEG analysis is primarily based on visual inspection, which is inherently subjective, operator-dependent, and difficult to standardize.

To address these limitations, quantitative EEG methods have been developed to provide objective and reproducible measures of neural activity [25, 30]. In particular, spectral approaches based on the Power Spectral Density (PSD), typically estimated using the Fourier transform, have become a standard tool for summarizing the distribution of signal energy across frequency bands. Clinical EEG interpretation, although based on standardized visual patterns, benefits from these quantitative tools, which improve objectivity and efficiency in demanding clinical environments [10, 19]. However, PSD-based methods rely on implicit stationarity assumptions and provide only a global frequency characterization, limiting their ability to capture transient and time-varying neural dynamics.

Recent advances in EEG signal processing combined with machine learning approaches have demonstrated promising performance in tasks such as epilepsy detection and classification [9, 15]. These methods provide a flexible framework for feature extraction, although challenges in interpretability remain [2, 17]. While data-driven approaches often achieve high predictive performance, their adoption in clinical practice is limited by the difficulty of relating extracted features to physiologically meaningful patterns [13].

In this context, symbolic and entropy-based representations have emerged as promising alternatives for describing the complexity of physiological signals. In particular, permutation-based approaches encode relationships between signal components through ordinal patterns, linking signal structure with information-theoretic measures [1, 31]. Symbolic representations have been shown to capture physiologically meaningful EEG dynamics [16], and similar entropy-based approaches have demonstrated strong discriminative power, particularly in epilepsy detection [22, 32]. However, most existing approaches operate directly on raw signals or fixed spectral features, without explicitly modeling the relative organization of energy across clinically relevant frequency bands.

The Weed Plot framework was previously introduced [6] as a method designed to bridge clinical electroencephalography (EEG) interpretation and quantitative analysis. This framework encodes band-wise spectral energy, typically obtained from Power Spectral Density via the Fast Fourier Transform (FFT), into a structured representation that preserves the interpretability of traditional EEG bands (Delta, Theta, Alpha, Beta1, Beta2, and Gamma). Classical PSD estimators, such as Welch’s method, provide a robust foundation for this representation [35].In particular, the Weed Plot framework captures the relative ranking of band energies, transforming spectral information into symbolic patterns that describe the dominant organization of the signal.

However, the original Weed Plot formulation relies on Fourier-based spectral estimation, which assumes signal stationarity and may fail to capture transient and non-stationary neural dynamics. EEG signals are inherently non-stationary, and their relevant features often evolve over time [21]. The limitations of purely spectral analysis motivate the use of time–frequency methods, where multi-resolution wavelet analysis provides both theoretical and practical advantages [4, 23]. Additionally, EEG recordings are often affected by artifacts, further emphasizing the need for robust representations capable of handling non-ideal conditions [34].

To address these limitations, time–frequency approaches based on the Discrete Wavelet Transform (DWT) have been widely adopted in EEG analysis. The DWT provides a multiresolution representation that enables simultaneous localization in time and frequency, making it particularly suitable for capturing transient neural activity [36, 37]. Wavelet-based methods have been successfully applied in feature extraction, denoising, and classification tasks due to their adaptability to non-stationary signals. In epileptic scenarios, recent studies have shown that wavelet-based decompositions combined with entropy measures effectively capture relevant dynamics and support competitive classification performance [12].

In this work, we propose a wavelet-based extension of the symbolic Weed Plot framework, where spectral energy is estimated using the DWT instead of traditional PSD methods. In contrast to approaches that directly associate wavelet decomposition levels with frequency bands, we define EEG bands a priori according to standard clinical ranges and use the wavelet transform to estimate band-wise energy within a time–frequency framework. This formulation preserves interpretability while incorporating temporal variability into the spectral representation. Entropy-based criteria are then applied to the resulting symbolic patterns to extract discriminative features [3].

The primary objective of this study is to evaluate the performance of this wavelet-based representation in comparison with both the original Fourier-based approach and raw signal analysis. Entropy features derived from these representations are assessed using multiple machine learning classifiers for neuro-disorder classification, using epilepsy as a benchmark case. The evaluation of such methods on clinically relevant EEG datasets is essential to ensure robustness and reproducibility in real-world scenarios [14, 11, 8, 27].

Our results show that the wavelet-based approach achieves comparable or improved classification performance while maintaining clinical interpretability, supporting the Weed Plot as a robust bridge between traditional EEG interpretation and automated diagnostic systems.

## 2. Preliminaries

### 2.1. Discrete EEG Signal and Frequency Bands

The brain generates a continuous-time analog signal *X*_*a*_(*t*). In digital electroencephalography (EEG), this signal is sampled at regular intervals *T*_*s*_ to produce a discrete sequence *x*(*n*) = *X*_*a*_(*nT*_*s*_), where *f*_*s*_ = 1/*T*_*s*_ is the sampling frequency.

Since EEG signals are inherently non-stationary, they are typically analyzed over short-time segments to capture the temporal evolution of neural dynamics. A fundamental aspect of this analysis is the decomposition of the signal into standard frequency bands. In clinical practice, these bands are defined as: *Delta* (0.5–4 Hz), *Theta* (4–8 Hz), *Alpha* (8–13 Hz), *Beta 1* (13–18 Hz), *Beta 2* (18–30 Hz), and *Gamma* (> 30 Hz). These ranges reflect different physiological states and constitute the basis of visual EEG interpretation (vEEG).For mathematical convenience, each frequency band is denoted as an interval *I*_*b*_, with *b* = 1, …, 6, where *I*_*b*_ corresponds to the frequency range associated with each clinical band.

### 2.2. Signal Representations: Fourier vs. Wavelets

To quantify how signal energy is distributed across these clinically relevant bands, it is necessary to adopt an appropriate signal representation. In this work, we consider two complementary mathematical frameworks.

In the frequency domain, the Discrete Fourier Transform (DFT) is employed:

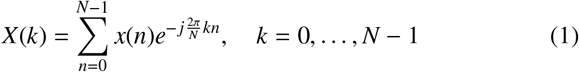

From this representation, the Power Spectral Density (PSD) is defined as *PS D*(*k*) = |*X*(*k*) |^2^, which provides a global description of how energy is distributed over frequencies. While the PSD is effective for stationary signals, it lacks temporal localization, which limits its ability to capture transient neural events. To overcome this limitation, a time–frequency representation can be used. The Discrete Wavelet Transform (DWT) provides a time–frequency representation of the signal by decomposing it into components at different scales. Unlike Fourier-based methods, which describe the global frequency content, the DWT captures localized variations, making it suitable for non-stationary signals such as EEG.

In practice, the DWT is implemented using a filter-bank structure that recursively separates the signal into approximation and detail coefficients. The approximation coefficients represent the low-frequency behavior of the signal, while the detail coefficients capture higher-frequency components at different scales.

In this work, the wavelet transform is used to estimate the energy distribution of the signal within predefined EEG frequency bands. For each band, the energy is computed from the corresponding wavelet coefficients as:

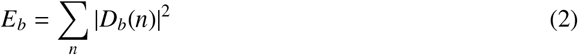

This formulation incorporates temporal variability into the spectral representation, allowing transient changes in the signal to be reflected in the estimated band energies, while preserving consistency with clinically defined frequency bands.

### 2.3. The Weed Plot Framework

Based on these signal representations, the next step is to quantify how the signal energy is distributed across clinically relevant frequency bands. In this context, the *Weed Plot* framework provides a formalization of the physician’s visual interpretation of EEG recordings by encoding the band-wise energy distribution into a structured feature vector.

In the Fourier-based approach, the *Accumulated Power per Band* (*AP*_*b*_) is defined as:

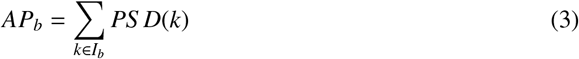

This leads to the vector 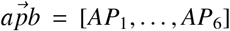, referred to as the *Accumulated Power per Band Vector*, which compactly summarizes the relative energy contribution of each frequency band and serves as the basis for subsequent symbolic and probabilistic representations.

In our proposed wavelet-based extension, the signal is decomposed using the Discrete Wavelet Transform (DWT) into approximation and detail coefficients. The resulting coefficients are used to estimate the energy distribution within each predefined EEG frequency band, rather than defining the bands themselves. More precisely, the energy of each band is computed as:

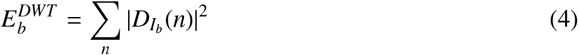

resulting in the vector 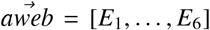, referred to as the *Accumulated Wavelet Energy per Band (AWEB) vector*, which captures both frequency-specific content and transient temporal dynamics. In contrast to the Fourier-based representation, this formulation provides a richer description of EEG activity by preserving time–frequency information.[24, 5]

## 3. The Weed Plot Quantification Method

While the vectors 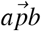 and 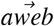 provide quantitative descriptions of energy distribution across frequency bands, clinical interpretation is primarily based on the relative dominance of these bands rather than their absolute values.

To bridge this gap, the proposed method formalizes the visual inspection of EEG recordings by mapping band-wise energy representations into a discrete symbolic space. This is achieved by encoding the relative ordering of frequency bands into symbolic patterns, capturing the hierarchical organization of energy across bands.

Unlike the original Weed Plot approach, where features are derived directly from the frequency of occurrence of symbolic patterns, we adopt a fully probabilistic formulation based on their empirical distribution. Specifically, each EEG segment is represented through the normalized frequency of band-ordering patterns, enabling a compact and statistically grounded characterization of brain dynamics.

### 3.1. Signal Segmentation and Symbolic Encoding

Given an EEG recording acquired under a specific montage with *C* channels, each channel *c* is analyzed independently. To capture the temporal evolution of the signal, a sliding window approach is employed, using windows of length Δ and step size τ, resulting in an overlap of Δ −τ. For each channel and each window *k*, the following procedure is applied:

Given an EEG recording acquired under a specific montage with *C* channels, each channel *c* is analyzed independently. To capture the temporal evolution of the signal, a sliding window approach is employed, using windows of length Δ and step size τ, resulting in an overlap of Δ −τ. For each channel and each window *k*, the following procedure is applied:

1. **Energy Quantification:** Compute the accumulated energy across the six standard clinical bands (*I*_1_, …, *I*_6_) using either the Fourier-based 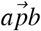 or the wavelet-based 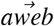 representations (see Section 2).
2. **Ranking:** Sort the frequency bands in decreasing order of energy to obtain a rank vector, where the highest-energy band is assigned rank 1 and the lowest-energy band rank 6. Each symbolic word can be naturally interpreted as an element of the symmetric group *S* _6_, representing a permutation of the six EEG frequency bands according to their relative energy. The *i*-th entry of 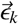 indicates the position of the *i*-th band in the ordered sequence.

This procedure transforms the continuous EEG signal of channel *c* into a sequence of symbolic words:

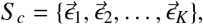

where *K* is the total number of sliding windows for that channel.

### 3.2. Channel-wise Probability Vectors

From the temporal sequence of symbolic words in each channel, we construct a *probability vector* **V**_*c*_ representing the empirical distribution over the vocabulary 𝒱:

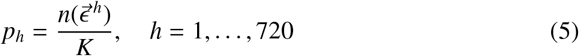

where 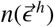 denotes the number of occurrences of the *h*-th word (according to lexicographic ordering), and *K* is the total number of sliding windows.

The resulting vector **V**_*c*_ = (*p*_1_, …, *p*_720_) defines a discrete probability distribution that provides a compact and robust signature of the spectral–temporal dynamics of channel *c*, inherently invariant to absolute amplitude scaling.

This probabilistic representation constitutes the basis of the *Weed Plot*, where each component *p*_*h*_ encodes the prevalence of a specific band-ordering pattern. The Weed Plot can thus be interpreted as a visualization of the empirical distribution of symbolic dynamics, highlighting dominant spectral organizations across time. Figure 1 illustrates the Weed Plot representations obtained from different signal quantification methods. While the PSD-based representation exhibits a relatively dispersed distribution across the vocabulary, the wavelet-based approaches (db4, sym4, coif5) produce more concentrated probability profiles, with dominant patterns localized in specific regions. This indicates that time–frequency representations yield more structured and discriminative symbolic signatures, potentially enhancing the separability of EEG dynamics in subsequent classification tasks.

**Figure 1:**
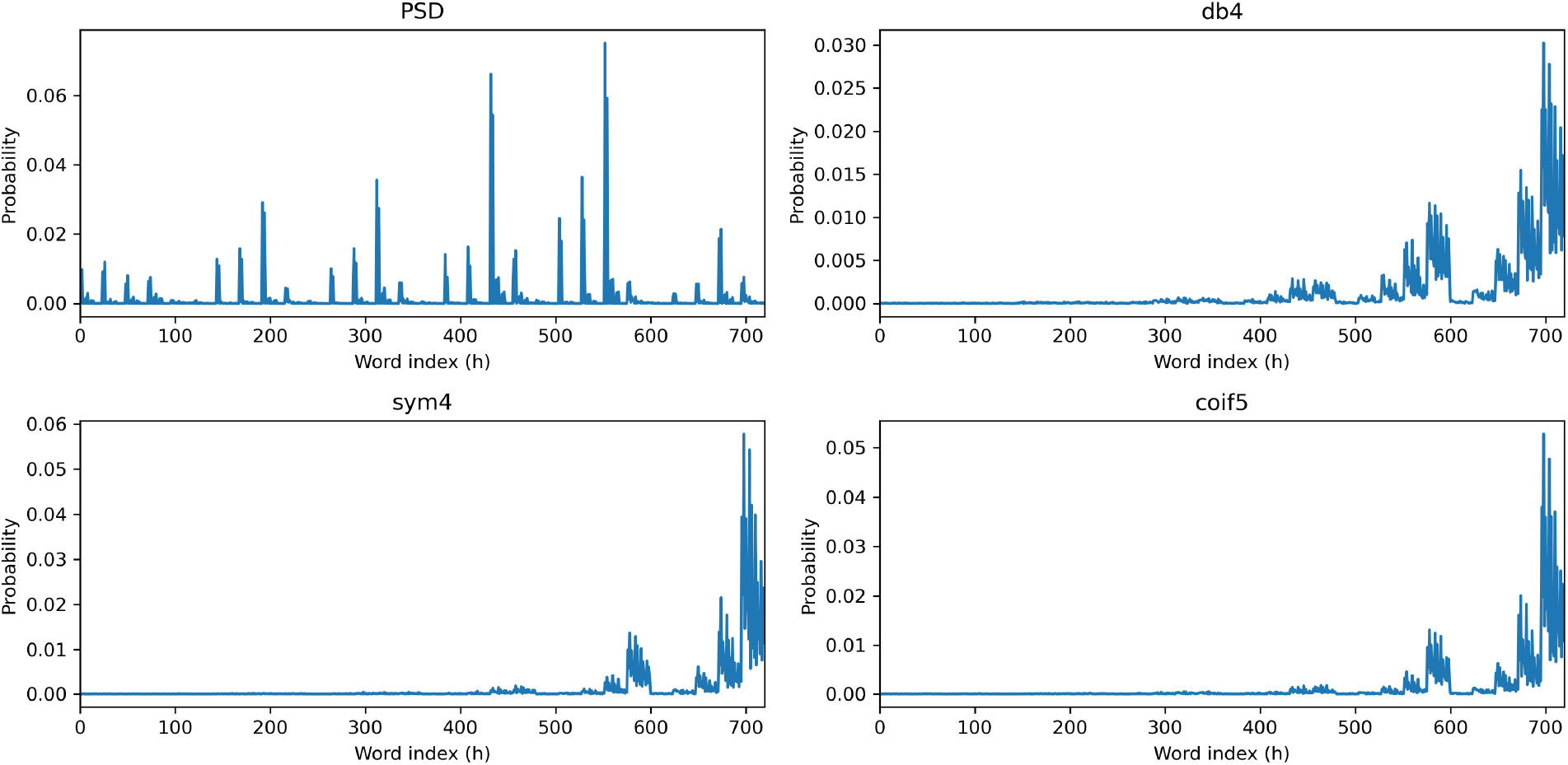
Comparison of Weed Plot probability distributions obtained using different signal representations: Power Spectral Density (PSD) and wavelet-based methods (db4, sym4, coif5). Each subplot shows the empirical probability of band-ordering patterns across the vocabulary of size 720.

### 3.3. Machine Learning Models

To assess the discriminative capability of the features derived from the proposed framework, a diverse set of machine learning models representing different learning paradigms was selected. This ensemble includes linear models such as Logistic Regression, margin-based techniques like Support Vector Machines (SVM), and ensemble methods based on bagging (Random Forest) and boosting (AdaBoost and Gradient Boosting). The inclusion of these varied architectures allows for a comprehensive evaluation of how different inductive biases interact with the symbolic spectral representations, determining whether class separability relies on linear decision boundaries or requires capturing the complex, non-linear interactions inherent in EEG dynamics.

Our main contributions through this work are summarized as follows:

- We propose a wavelet-based extension of the Weed Plot framework, improving the analysis of non-stationary EEG signals by leveraging the superior time–frequency localization of the Discrete Wavelet Transform (DWT).
- We provide a comprehensive evaluation of symbolic spectral features across diverse learning paradigms, including linear baselines (LR), margin-based methods (SVM), and ensemble architectures (Random Forest, AdaBoost, and Gradient Boosting).
- We conduct a comparative analysis between Fourier-based and wavelet-based energy quantification, demonstrating that the interaction between symbolic encoding and wavelet decomposition yields more discriminative features for epilepsy detection than traditional spectral methods or raw signal descriptors.

## 4. Materials and Methods

This section describes the experimental framework, subject selection criteria, and the preprocessing pipeline designed to standardize EEG signals for classification (see Fig. 2).

**Figure 2:**
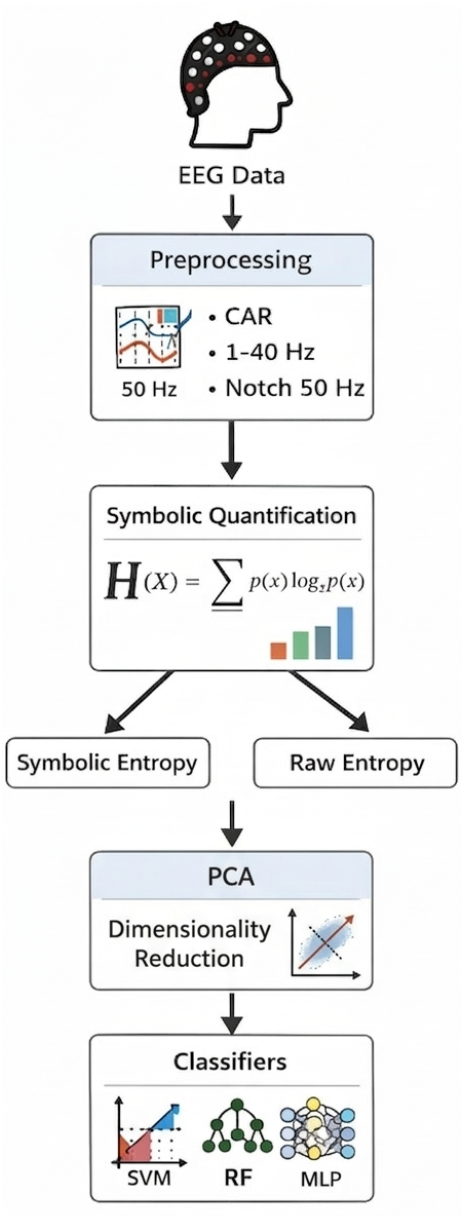
Overview of the proposed EEG processing pipeline.

### 4.1. Data Sources and Subject Selection

To evaluate the proposed quantification methods, we combined two open-source datasets to compare healthy baseline activity against pathological epileptic dynamics.

The pathological group was sourced from the Temple University Hospital (TUH) EEG Epilepsy Corpus (TUEP) v3.0.0 [28]. To ensure data quality and minimize pharmacological confounds, we excluded subjects under anti-epileptic treatment. The final cohort consisted of 26 unmedicated subjects exhibiting clinical or electrographic seizures, with recordings exceeding 15 minutes to ensure statistical robustness. Both interictal and ictal segments were included to capture non-stationary dynamics.

The control group comprises 14 healthy subjects from a public dataset [20], featuring resting-state, eyes-closed EEG recordings without neurological disorders or medication, providing a baseline for normal brain activity.

### 4.2. Preprocessing and Digital Standardization

EEG recordings were processed using the *MNE-Python* library [18]. Signals were resampled to 250 Hz and re-referenced using a Common Average Reference (CAR) scheme. To ensure consistency, electrode labels were mapped to the international 10–20 system.

The cleaning pipeline included a notch filter at 50 Hz to remove power-line interference, followed by a 4th-order Butterworth band-pass filter (1–40 Hz) to suppress artifacts. Finally, the standardized signals were decomposed into the six clinical frequency bands (*I*_1_, …, *I*_6_) as defined in Section 2. To capture non-stationary dynamics, the first 15 minutes of each EEG recording were segmented into 1-s windows with 50% overlap (τ = 0.5 s). This configuration yields approximately 1800 windows per channel, balancing temporal resolution and statistical robustness.

For each window, energy distribution across frequency bands was estimated using two approaches:

- *Fourier-based:* Power Spectral Density (PSD) was computed via Welch’s method, averaging modified periodograms.
- *Wavelet-based:* Discrete Wavelet Transform (DWT) was applied using *db4, sym4*, and *coif5* kernels, selected for their superior time–frequency localization in EEG analysis.

Based on the band-wise energy, a symbolic representation was constructed by ranking the six frequency bands in decreasing order. This produces a permutation vector (symbolic *word*) for each window. With six bands, the vocabulary consists of 6! = 720 possible states, encoding the relative dominance of spectral content. The complexity of the spectral organization was quantified using Shannon entropy [31]:

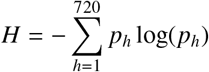

where *p*_*h*_ is the probability of the *h*-th symbolic word. As a time-domain baseline, the normalized Permutation Entropy [1] (*m* = 5) was computed directly from the preprocessed signals. This measure quantifies the diversity of ordinal patterns, allowing us to compare the intrinsic temporal complexity of the EEG against the proposed symbolic spectral dynamics.

### 4.3. Statistical Analysis of Entropy Distributions

To evaluate group differences, we compared entropy values across 19 EEG channels using boxplot visualizations (Fig. 3) and the non-parametric *Mann–Whitney U-test*. The latter was chosen due to the non-Gaussian distribution of the entropy data and the presence of outliers.

**Figure 3:**
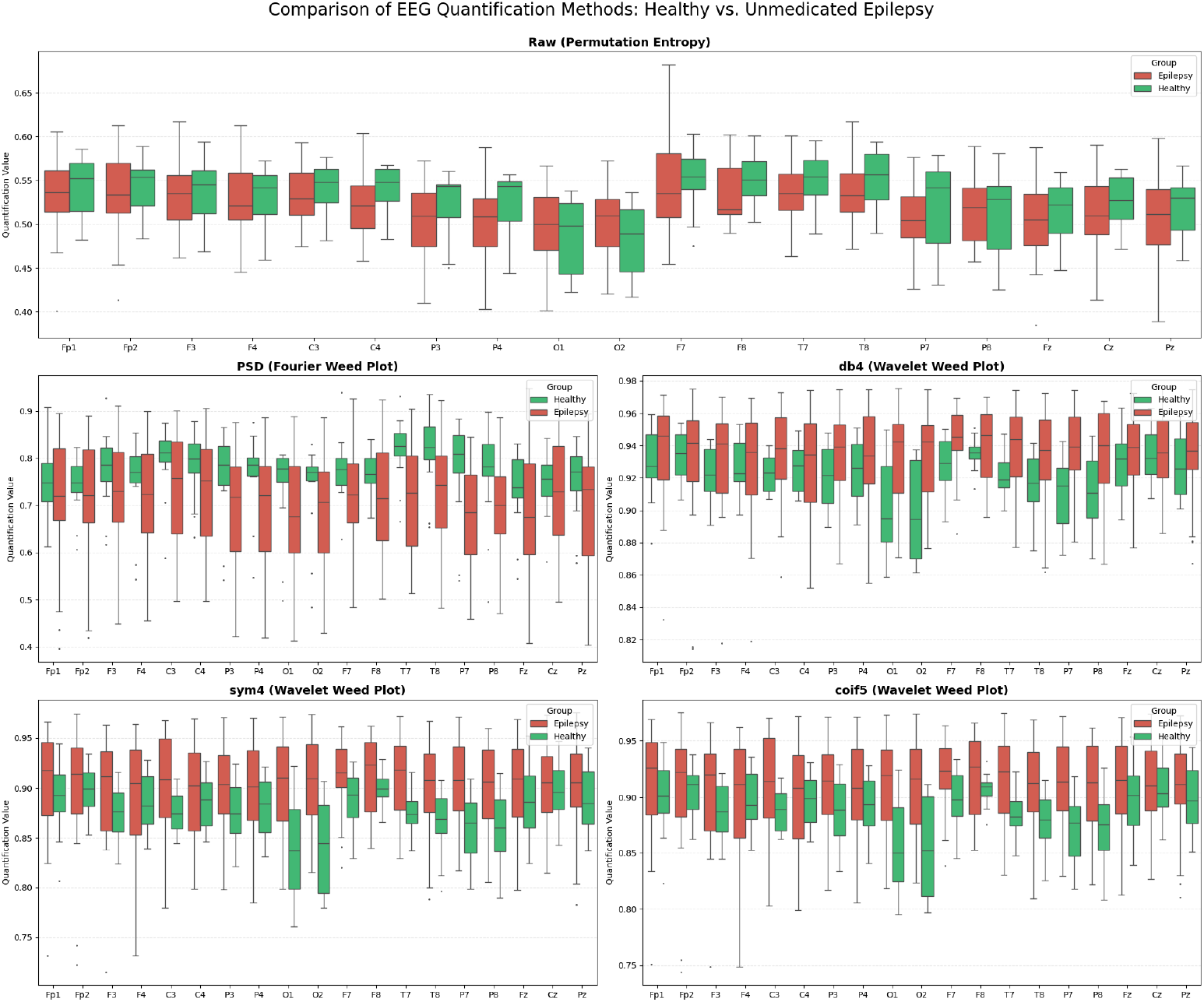
Distribution of entropy values across 19 EEG channels for control and epileptic groups. Subplots compare raw signal permutation entropy against the proposed symbolic spectral entropy.

As summarized in Table 1, raw signal permutation entropy failed to show statistically significant differences (*p* > 0.05) in most channels. In contrast, the proposed symbolic spectral entropy—specifically using wavelet-based representations—revealed significant discriminative patterns (*p* < 0.05) in temporal (*T* 7, *T* 8), parietal (*P*7, *P*8), and occipital (*O*1, *O*2) regions. This suggests that the symbolic encoding of spectral dynamics provides a more robust representation of epileptic activity than time-domain complexity measures.

**Table 1:** Statistical significance comparison (Mann-Whitney U-test) across EEG channels. *p* < 0.05 (indicated as YES) denotes a significant difference between groups.

| Channel | Raw |  | PSD |  | db4 |  | sym4 |  | coif5 |  |
| --- | --- | --- | --- | --- | --- | --- | --- | --- | --- | --- |
|  | p-val | Sig. | p-val | Sig. | p-val | Sig. | p-val | Sig. | p-val | Sig. |
| Fp1 | 0.7019 | no | 0.5999 | no | 0.2281 | no | 0.1873 | no | 0.1970 | no |
| Fp2 | 0.7659 | no | 0.5803 | no | 0.3871 | no | 0.3007 | no | 0.2627 | no |
| F3 | 0.9661 | no | 0.2393 | no | 0.0812 | no | **0.0371** | YES | **0.0456** | YES |
| F4 | 0.7230 | no | 0.5235 | no | 0.4190 | no | 0.3141 | no | 0.3421 | no |
| C3 | 0.3717 | no | 0.1690 | no | 0.0862 | no | **0.0488** | YES | 0.0593 | no |
| C4 | 0.0718 | no | 0.1690 | no | 0.4696 | no | 0.4355 | no | 0.4872 | no |
| P3 | 0.0916 | no | 0.0718 | no | 0.1366 | no | 0.0812 | no | 0.1030 | no |
| P4 | 0.0593 | no | **0.0398** | YES | 0.5610 | no | 0.3871 | no | 0.4524 | no |
| O1 | 0.3007 | no | 0.1155 | no | **0.0030** | YES | **0.0019** | YES | **0.0012** | YES |
| O2 | 0.1970 | no | 0.1366 | no | **0.0008** | YES | **0.0008** | YES | **0.0007** | YES |
| F7 | 0.6197 | no | 0.0633 | no | 0.0674 | no | **0.0301** | YES | **0.0398** | YES |
| F8 | 0.1222 | no | 0.1442 | no | 0.2627 | no | 0.2174 | no | 0.1970 | no |
| T7 | 0.1155 | no | **0.0131** | YES | **0.0131** | YES | **0.0074** | YES | **0.0068** | YES |
| T8 | 0.2070 | no | **0.0052** | YES | **0.0301** | YES | **0.0179** | YES | **0.0179** | YES |
| P7 | 0.1873 | no | **0.0068** | YES | **0.0068** | YES | **0.0052** | YES | **0.0052** | YES |
| P8 | 0.8316 | no | **0.0301** | YES | **0.0142** | YES | **0.0074** | YES | **0.0087** | YES |
| Fz | 0.8316 | no | 0.2750 | no | 0.1780 | no | 0.1970 | no | 0.2070 | no |
| Cz | 0.2750 | no | 0.7876 | no | 0.7659 | no | 0.6810 | no | 0.6399 | no |
| Pz | 0.4355 | no | 0.2393 | no | 0.3421 | no | 0.3421 | no | 0.3279 | no |

### 4.4. Feature Space Analysis and Dimensionality Reduction

To mitigate spatial redundancy across the 19 EEG channels, Principal Component Analysis (PCA) was applied after standardizing the feature vectors (*S tandardS caler* [29]). As illustrated in Fig. 4, we retained the principal components (PCs) required to explain 95% of the total variance. Notably, wavelet-based methods (*sym4, coif5*) exhibited faster variance convergence compared to raw permutation entropy, suggesting a more compact representation of pathological dynamics.

**Figure 4:**
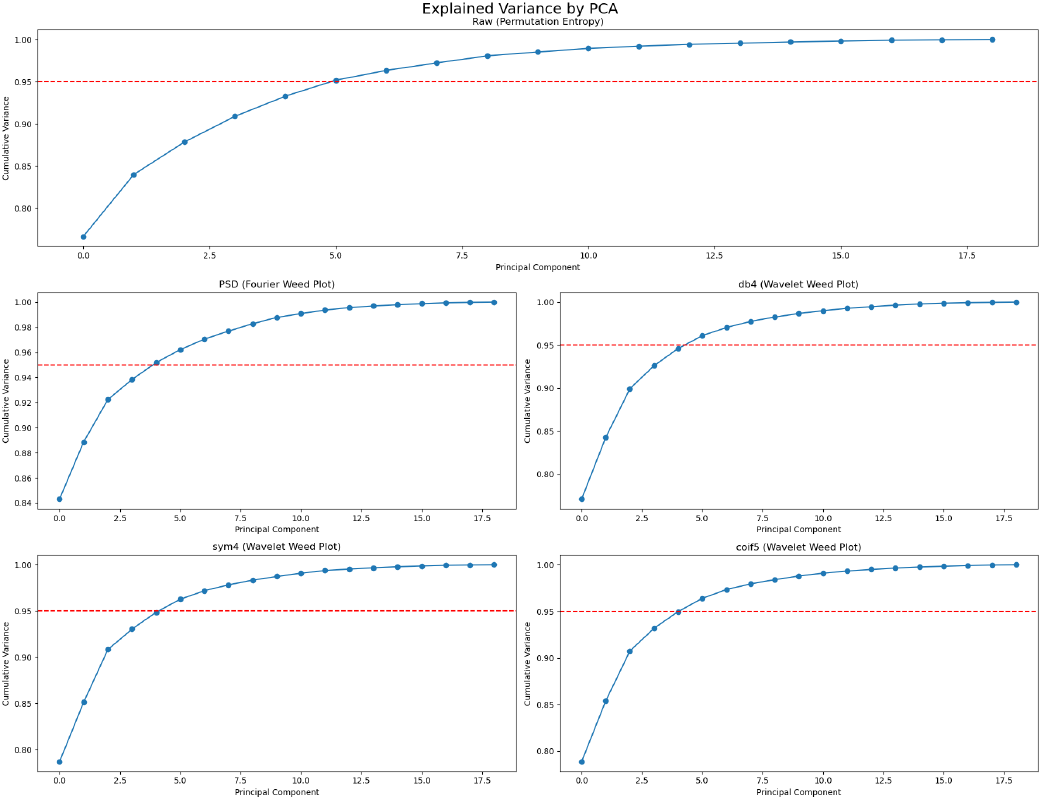
Cumulative explained variance and 95% threshold (red dashed line) for each quantification method.

The topographic analysis of PCA loadings (Fig. 5) indicates that while PC1 represents a global complexity shift across all electrodes, PC2 captures regional specialization. In the wavelet-based Weed Plots, PC2 emphasizes frontal (*Fp*1, *Fp*2) and parieto-occipital regions, aligning with the significant channels identified in the Mann–Whitney analysis. Finally, the three dimensional projection of the first three PCs (Fig. 6) visually confirms that symbolic spectral encoding enhances group segregation compared to the raw signal baseline, providing a more separable input space for the subsequent machine learning stage.

**Figure 5:**
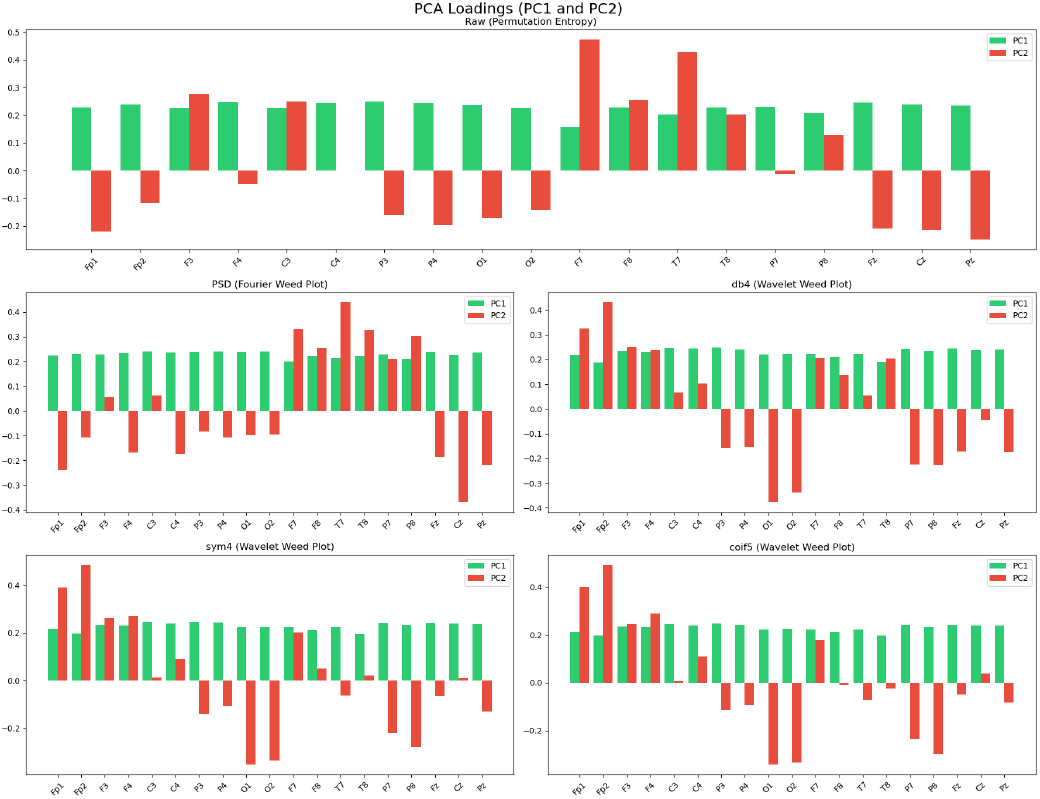
PCA loadings for the first two principal components (PC1 and PC2) across all EEG channels and quantification methods. The bar height represents the contribution of each electrode to the corresponding principal component.

**Figure 6:**
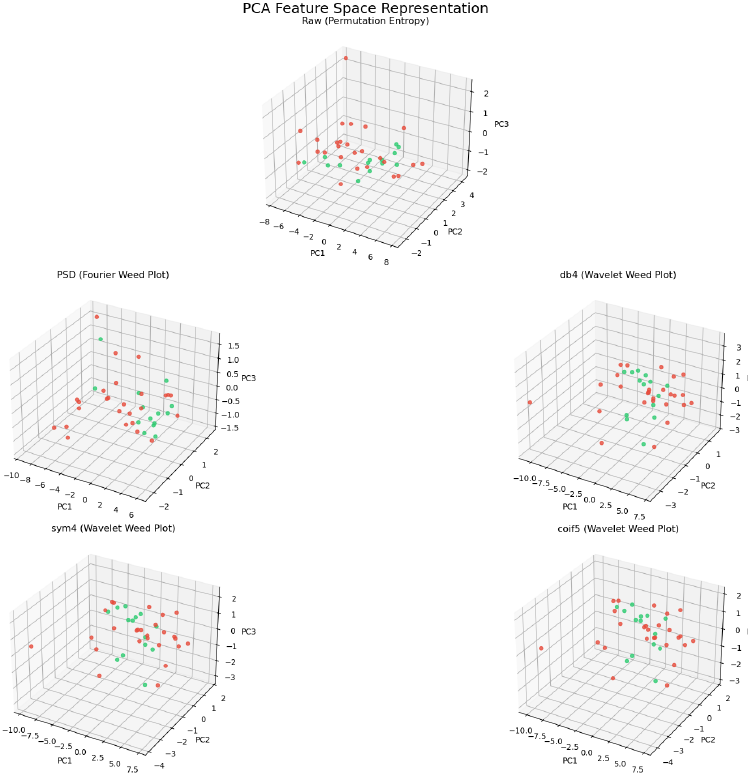
Three–dimensional PCA projection of the entropy feature vectors using the first three principal components. Green points correspond to control subjects, while red points represent epileptic subjects.

**Figure 7:**
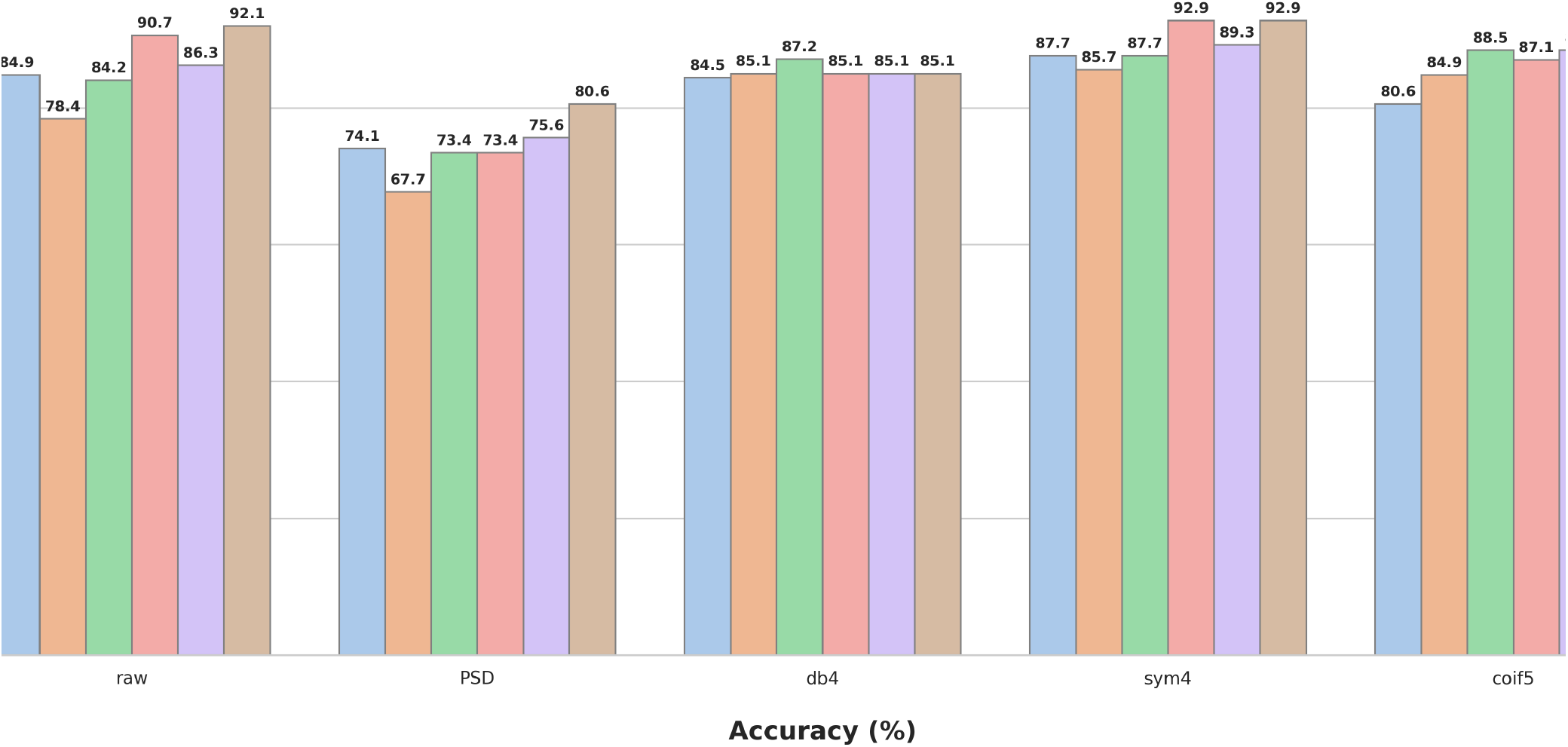
* Figure 4: ACCURACY (%) of six Classifiers LR, SVM, DT, RF, GB, AB.

**Figure 8:**
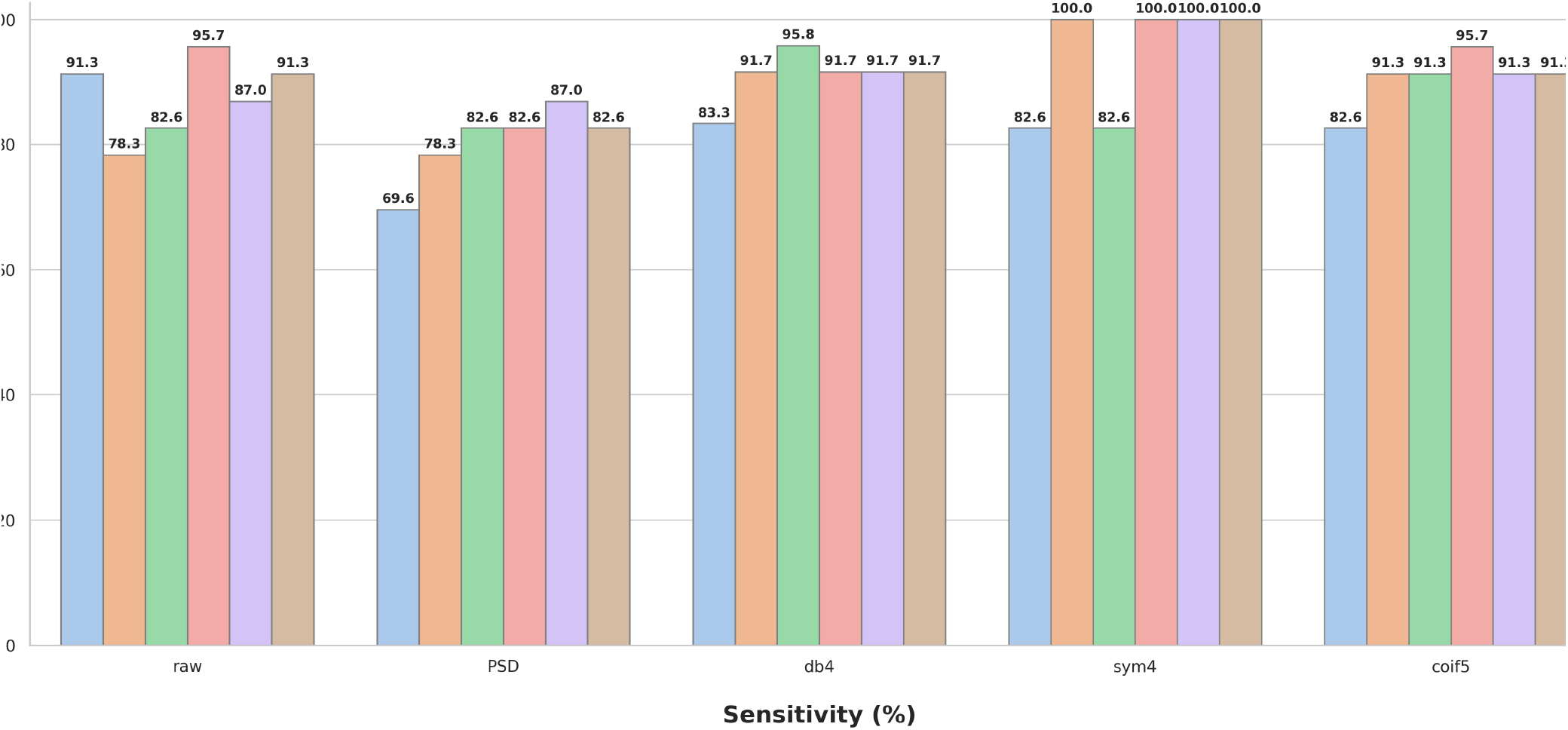
* Figure 5: SENSITIVITY (%) of six Classifiers LR, SVM, DT, RF, GB, AB.

**Figure 9:**
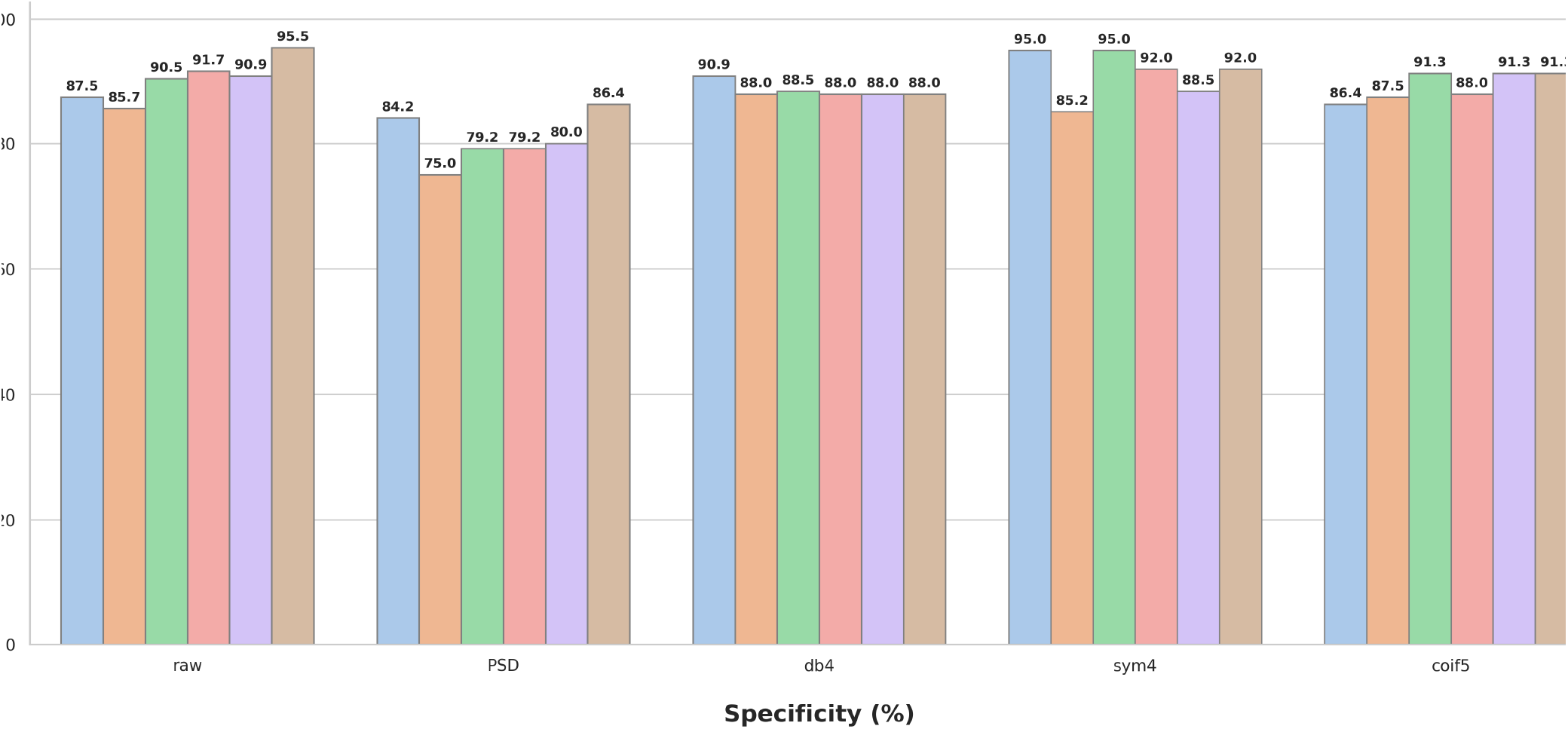
* Figure 6: SPECIFICITY (%) of six Classifiers LR, SVM, DT, RF, GB, AB.

**Figure 10:**
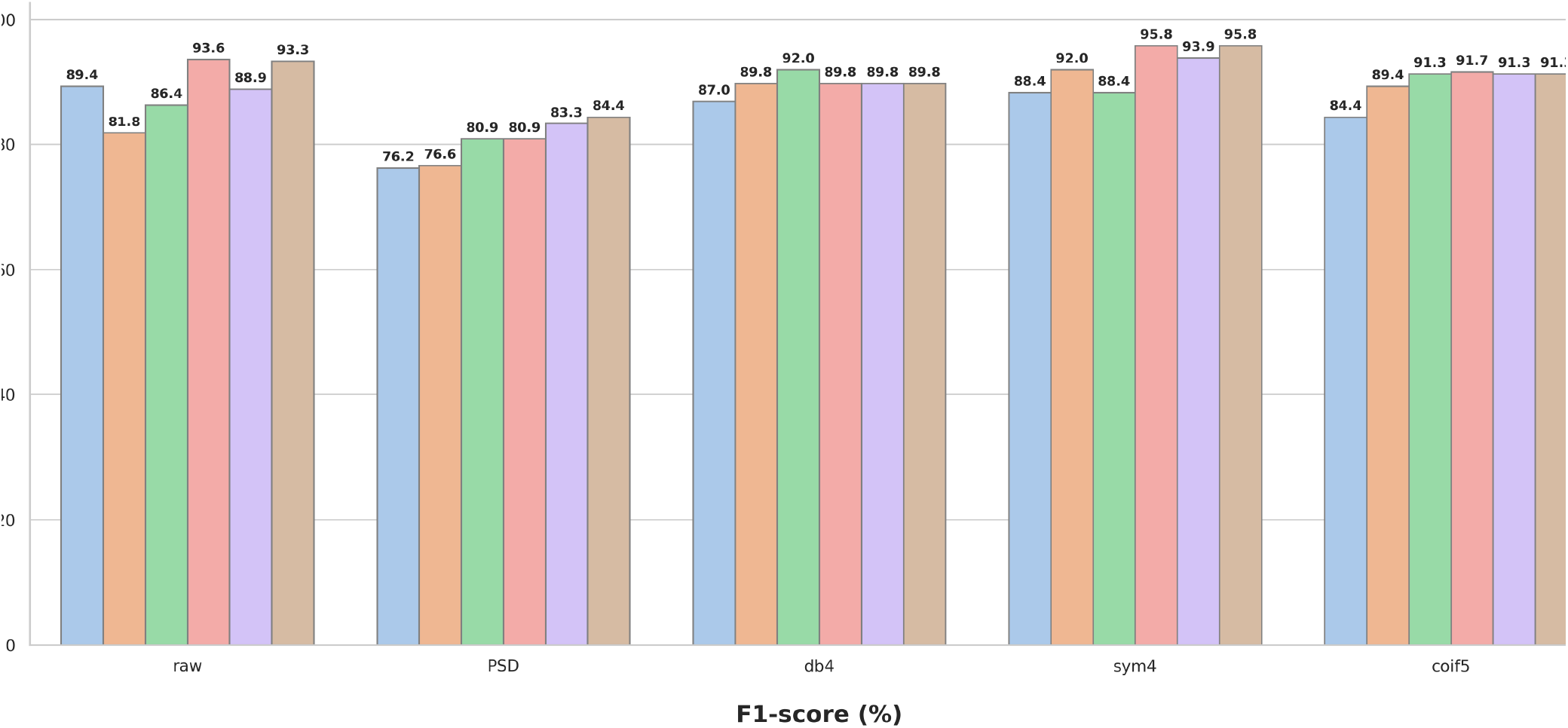
* Figure 7: F1-SCORE (%) of six Classifiers LR, SVM, DT, RF, GB, AB.

### 4.5. Classification Framework

#### 4.5.1. Model Construction and Hyperparameter Tuning

To ensure a fair comparison across the different quantification methods (Raw, PSD, and Wavelet Weed Plots), hyperparameter optimization was performed independently for each combination of model and feature set. A *grid search* strategy was employed within a *nested cross-validation framework*. Specifically, for each fold of the outer 5-fold cross-validation, an inner validation loop was used to select the optimal hyperparameters.

To guarantee reproducibility and consistency across experiments, a fixed random seed (*random*_*state* = 42) was used for all stochastic processes, including data splitting, PCA initialization, and model training.

Tables 2–6 summarize the optimal hyperparameter configurations obtained.

**Table 2:**
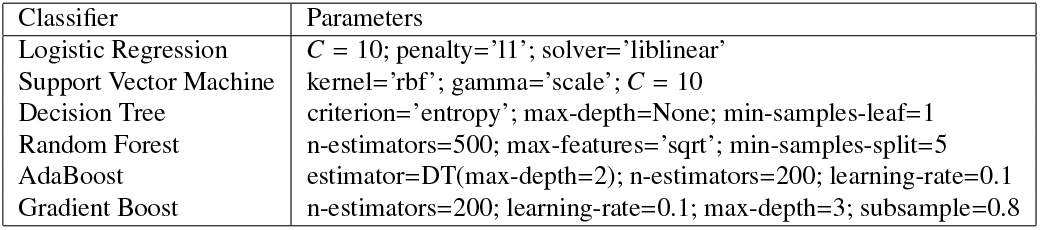
Hyperparameters for Raw Permutation Entropy.

**Table 3:** Hyperparameters for PSD (Fourier Weed Plot).

| Classifier | Parameters |
| --- | --- |
| Logistic Regression | $C = 1$; penalty='l2'; solver='liblinear' |
| Support Vector Machine | kernel='rbf'; gamma='auto'; $C = 1$ |
| Decision Tree | criterion='gini'; min-samples-leaf=2 |
| Random Forest | n-estimators=200; max-features='sqrt'; min-samples-split=2 |
| AdaBoost | estimator=DT(max-depth=2); n-estimators=200; learning-rate=0.1 |
| Gradient Boost | n-estimators=100; learning-rate=0.01; max-depth=2; subsample=1.0 |

**Table 4:** Hyperparameters for db4 (Wavelet Weed Plot).

| Classifier | Parameters |
| --- | --- |
| Logistic Regression | $C = 100$; penalty='l2'; solver='liblinear' |
| Support Vector Machine | kernel='rbf'; gamma='scale'; $C = 1$ |
| Decision Tree | criterion='entropy'; min-samples-leaf=3 |
| Random Forest | n-estimators=200; max-features='sqrt'; min-samples-split=2 |
| AdaBoost | estimator=DT(max-depth=2); n-estimators=50; learning-rate=0.1 |
| Gradient Boost | n-estimators=200; learning-rate=0.01; max-depth=2; subsample=0.8 |

**Table 5:** Hyperparameters for sym4 (Wavelet Weed Plot).

| Classifier | Parameters |
| --- | --- |
| Logistic Regression | $C = 10$; penalty='l1'; solver='liblinear' |
| Support Vector Machine | kernel='rbf'; gamma='scale'; $C = 10$ |
| Decision Tree | criterion='entropy'; min-samples-leaf=3 |
| Random Forest | n-estimators=800; max-features='sqrt'; min-samples-split=2 |
| AdaBoost | estimator=DT(max-depth=2); n-estimators=50; learning-rate=0.05 |
| Gradient Boost | n-estimators=100; learning-rate=0.1; max-depth=3; subsample=1.0 |

**Table 6:**
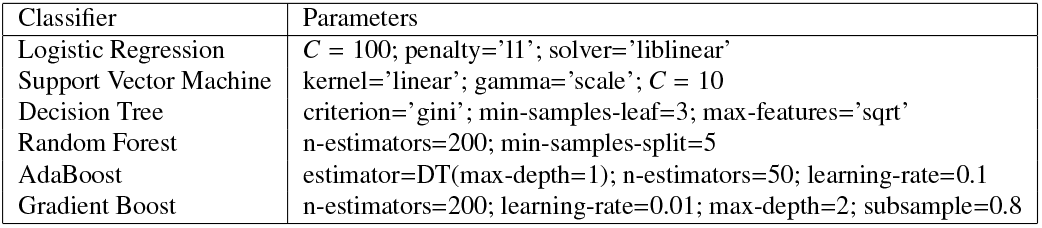
Hyperparameters for coif5 (Wavelet Weed Plot).

### 4.6. Performance Evaluation

Due to the class imbalance (26 epileptic vs. 14 control subjects), classifier performance was assessed using balanced metrics to ensure a reliable evaluation. We employed *Balanced Accuracy* (the average of sensitivity and specificity), *F1-score*, and the *Area Under the Receiver Operating Characteristic Curve (AUC)*. While the F1-score provides a robust measure of the precision-recall balance, the AUC offers a threshold-independent assessment of the models’ discriminative power. These metrics were selected to prioritize the correct identification of pathological dynamics while maintaining high specificity in the healthy control group.

## 5. Results

The classification performance across the six machine learning models and five feature representations is summarized in Table 7. Overall, wavelet-based Weed Plot quantifiers consistently outperformed both raw permutation entropy and PSD-based features across all evaluation metrics. This superior performance aligns with the higher variability in band-ordering patterns induced by wavelet decomposition, resulting in more discriminative symbolic distributions.

**Table 7:** Performance metrics (%) with optimized thresholds (outliers removed). Wavelet-based kernels (coif5, db4, sym4) are presented first, followed by PSD and Raw signal baselines.

| Model | Accuracy |  |  |  |  | Sensitivity (Recall) |  |  |  |  | Specificity |  |  |  |  | F1-score |  |  |  |  | AUC |  |  |  |  |
| --- | --- | --- | --- | --- | --- | --- | --- | --- | --- | --- | --- | --- | --- | --- | --- | --- | --- | --- | --- | --- | --- | --- | --- | --- | --- |
|  | coif5 | db4 | sym4 | PSD | Raw | coif5 | db4 | sym4 | PSD | Raw | coif5 | db4 | sym4 | PSD | Raw | coif5 | db4 | sym4 | PSD | Raw | coif5 | db4 | sym4 | PSD | Raw |
| LR | 81.0 | 85.0 | 89.0 | 84.0 | 85.0 | 83.0 | 83.0 | 100.0 | 91.0 | 85.0 | 86.0 | 91.0 | 86.0 | 83.0 | 91.0 | 84.0 | 87.0 | 93.0 | 87.0 | 88.0 | 89.4 | 90.2 | 93.2 | 90.9 | 89.6 |
| SVM | 85.0 | 85.0 | 82.0 | 77.0 | 84.0 | 91.0 | 91.0 | 81.0 | 73.0 | 88.0 | 88.0 | 88.0 | 88.0 | 83.0 | 85.0 | 89.0 | 90.0 | 84.0 | 78.0 | 87.0 | 88.2 | 92.3 | 82.1 | 71.4 | 79.7 |
| DT | 89.0 | 89.0 | 85.0 | 83.0 | 89.0 | 91.0 | 91.0 | 85.0 | 83.0 | 92.0 | 91.0 | 91.0 | 89.0 | 89.0 | 92.0 | 91.0 | 91.0 | 87.0 | 86.0 | 92.0 | 89.3 | 83.5 | 84.9 | 67.3 | 91.5 |
| RF | 87.0 | 85.0 | 82.0 | 72.0 | 83.0 | 96.0 | 92.0 | 85.0 | 73.0 | 92.0 | 88.0 | 88.0 | 88.0 | 83.0 | 91.0 | 92.0 | 90.0 | 86.0 | 78.0 | 91.0 | 89.9 | 89.0 | 86.8 | 71.4 | 86.5 |
| GB | 89.0 | 80.0 | 84.0 | 75.0 | 84.0 | 91.0 | 96.0 | 96.0 | 92.0 | 88.0 | 91.0 | 83.0 | 86.0 | 80.0 | 88.0 | 91.0 | 89.0 | 91.0 | 86.0 | 88.0 | 86.2 | 87.5 | 89.4 | 75.3 | 89.3 |
| AB | 89.0 | 86.0 | 86.0 | 50.0 | 84.0 | 91.0 | 100.0 | 100.0 | 100.0 | 88.0 | 91.0 | 87.0 | 87.0 | 65.0 | 88.0 | 91.0 | 93.0 | 93.0 | 79.0 | 88.0 | 89.4 | 96.4 | 89.8 | 31.6 | 88.5 |

Specifically, *Gradient Boosting* and *AdaBoost* achieved the highest balanced accuracy (89%) when using *coif5* kernel. In contrast, PSD-based representations showed a significant performance decay, with accuracies dropping to 50% in ensemble methods. Regarding sensitivity, several models (e.g., Logistic Regression and AdaBoost) reached 100% detection of epileptic subjects using wavelet features or PDS, demonstrating their ability to capture pathological signatures.

The *AUC* results further confirm this trend, with wavelet representations reaching 96.4% (*db4*) and 89.9% (*coif5*), indicating excellent class separability across decision thresholds. While raw entropy provides a baseline and PSD captures stationary spectral content, the integration of time–frequency information through wavelets significantly enhances the robustness of entropy-based descriptors for epilepsy classification.

## 6. Discussion

In this study, we evaluated the discriminative capability of entropy-based features derived from both raw EEG signals and symbolic spectral representations obtained through the Weed Plot framework. The results consistently demonstrate that the proposed symbolic encoding, particularly when combined with wavelet-based spectral quantification, enhances the separability between epileptic and control subjects.

A central finding of this work is that entropy computed over symbolic spectral representations exhibits significantly higher discriminative power than raw permutation entropy. While raw entropy captures temporal complexity at the signal level, it fails to fully characterize the hierarchical organization of frequency bands. In contrast, the Weed Plot representation encodes the relative dominance of spectral components, providing a structured and interpretable description of EEG dynamics aligned with clinical reasoning.

Among the evaluated spectral approaches, wavelet-based methods (db4, sym4, and coif5) consistently outperformed the Fourier-based PSD representation. Although both approaches ultimately estimate energy per frequency band, they differ fundamentally in how this energy is obtained. The PSD provides a global representation of frequency content, effectively averaging signal behavior over time. As a result, band energies tend to remain stable across temporal windows, leading to limited variability in the relative ordering of frequency bands.

In contrast, the Discrete Wavelet Transform (DWT) decomposes the signal using localized basis functions, producing coefficients that capture transient and time-dependent fluctuations in neural activity. While the final energy measure is obtained by aggregating wavelet coefficients, the intermediate representation preserves temporal localization. Consequently, the estimated band energies are sensitive to local signal dynamics, reflecting transient changes that are not captured by Fourier-based methods.

This distinction plays a crucial role in the symbolic encoding stage of the Weed Plot framework. Each temporal window is represented by a permutation that encodes the ranking of band energies. Therefore, the variability of these rankings across time directly determines the structure of the symbolic sequence. In the Fourier-based approach, the relative ordering of bands tends to remain stable, generating a limited set of recurring symbolic patterns and resulting in a more diffuse empirical distribution over the permutation space.

Conversely, the wavelet-based representation induces greater variability in band-ordering patterns due to its sensitivity to transient events. This leads to richer symbolic sequences, where specific permutations emerge as dominant under certain temporal conditions. As a result, the empirical probability distribution over the vocabulary becomes more structured, with higher concentration around characteristic patterns, thereby improving class separability.

From a machine learning perspective, models such as Support Vector Machines and ensemble methods (Random Forest and AdaBoost) achieved the best performance across most feature representations. However, beyond raw performance metrics, models trained on wavelet-derived features exhibited greater stability and a better balance between sensitivity and specificity, which is critical in clinical applications.

The use of Principal Component Analysis (PCA) further contributed to reducing redundancy across channels while preserving the most informative variability. Notably, the faster convergence of explained variance observed in wavelet-based features suggests that pathological information is more compactly represented in this domain.

Despite these promising results, some limitations should be acknowledged. The dataset size remains relatively small, particularly for the control group, which may affect the generalizability of the models. Additionally, while entropy provides a compact descriptor, it inevitably reduces the richness of the original signal, potentially discarding fine-grained temporal information.

Future work could formalize the symbolic representation as a stochastic process over permutations, enabling the analysis of transition dynamics, entropy rates, and divergence measures between classes. Additionally, extending this framework to larger datasets and integrating deep learning architectures could further enhance classification performance.

Overall, the findings suggest that the combination of symbolic spectral encoding and wavelet-based quantification provides a robust and interpretable framework for capturing the complex dynamics of EEG signals, offering clear advantages over traditional time-domain and Fourier-based approaches.

## 7. Conclusion

This study demonstrates that replacing Fourier-based spectral estimation with wavelet-based quantification significantly enhances the capture of non-stationary dynamics in EEG signals. The primary advantage of this approach lies in the synergy between wavelet time–frequency localization and symbolic encoding, which produces more informative feature structures that align with clinical interpretation. Our results confirm that wavelet-derived Weed Plot quantifiers provide superior discriminative power and class separability compared to traditional raw entropy or PSD-based baselines.

Future work will focus on validating this framework across larger, multi-center datasets and exploring temporal dependencies through stochastic modeling of symbolic transitions. Additionally, the topographic insights provided by PCA loadings suggest a promising path for the clinical localization of neural dynamics. Overall, the proposed combination of symbolic encoding and wavelet representations offers a robust, interpretable, and computationally efficient framework for EEG characterization, effectively bridging the gap between expert visual analysis and automated diagnostic tools.

## Declaration of Generative AI and AI-assisted technologies in the writing process

During the preparation of this work, the authors used Gemini (Google) in order to improve the language, grammatical correctness, and overall readability of the manuscript. After using this tool, the authors reviewed and edited the content as needed and take full responsibility for the final publication’s content.

## Data availability

The datasets analyzed in this study are available in public repositories. The pathological group data was sourced from the Temple University Hospital (TUH) EEG Epilepsy Corpus (TUEP) v3.0.0 [28]. The healthy control group data is available through the public dataset provided in [20].

